# Reproducible neuroimaging features for diagnosis of Autism spectrum disorder with machine learning

**DOI:** 10.1101/2021.10.21.21265162

**Authors:** Cooper J. Mellema, Kevin P. Nguyen, Alex Treacher, Albert Montillo

**Affiliations:** Lyda Hill Department of Bioinformatics, UT Southwestern; Biomedical Engineering Department, UT Southwestern; Advanced Imaging Research Center, UT Southwestern; Radiology Department, UT Southwestern

**Keywords:** Autism Spectrum Disorder, Diagnosis, Machine Learning, Neuroimaging, functional MRI, structural MRI, biomarkers

## Abstract

Autism spectrum disorder (ASD) is the fourth most common neurodevelopmental disorder, with a prevalence of 1 in 160 children. Accurate diagnosis relies on experts, but such individuals are scarce. This has led to increasing interest in the development of machine learning (ML) models that can integrate neuroimaging features from functional and structural MRI (fMRI and sMRI) to measure alterations manifest in ASD. We optimized and compared the performance of 12 of the most popular and powerful ML models. Each was separately trained using 15 different combinations of fMRI and sMRI features and optimized with an unbiased model search. Deep learning models predicted ASD with the highest diagnostic accuracy and generalized well to other MRI datasets. Our model achieves state-of-the-art 80% area under the ROC curve (AUROC) in diagnosis on test data from the IMPAC dataset; and 86% and 79% AUROC on the external ABIDE I and ABIDE II datasets. The highest performing models identified reproducible putative biomarkers for accurate ASD diagnosis in accord with known ASD markers as well as novel cerebellum biomarkers. Such reproducibility lends credence to their tremendous potential for defining and using a set of truly generalizable ASD biomarkers that will advance scientific understanding of neuronal changes in ASD.

## 1 Introduction

Autism Spectrum Disorder (ASD) is currently diagnosed through a time-consuming evaluation of behavioral tests by expert clinicians specializing in neurodevelopmental disorders. This diagnosis can be challenging due to several factors including the heterogeneity of the spectrum disorder, the uncertainty in the administration and interpretation of behavioral tests, and neurobiological and phenotypical differences that vary only slightly compared to typically developing controls [1]. These differences are believed to be due to altered neural connectivity in participants with ASD. A standardized and accurate diagnostic tool would increase availability and reproducibility of diagnostic services while reducing subjectivity [1–3]. As candidate noninvasive measures to facilitate diagnosis, functional MRI (fMRI) and structural MRI (sMRI) quantify brain connectivity and 3-dimensional structure, respectively. The blood oxygen level dependent (BOLD) signal from fMRI measures changes in flow and the ratio of oxy/deoxyhemoglobin in the blood throughout the brain; an indirect measure of neural activity. As ASD is putatively a neural connectivity disorder, regional signals can then be converted to interregional functional connectivity measures (FC). Structural MRI enables the quantification of complementary measures of brain morphology such as cortical thickness and subcortical structure volume.

Previous attempts to automate diagnosis through neuroimaging and machine learning are limited in three ways. First, they typically focus on one proposed predictive model and do not equally optimize or tune the hyperparameters of alternative methods, leading to biased results. Second, once a model is generated, they do not thoroughly analyze the model to reveal the learned biomarkers or discuss the neurophysiological significance of the findings. Third, they do not validate or adapt their models to an external dataset and are thereby prone to a spurious result applicable only to the single dataset used for model construction.

We conducted a systematic comparison of linear, nonlinear, and deep learning ML models and assessed their relative performances using a large ASD dataset. Hyperparameters of each model were carefully optimized to a similar degree to avoid preferentially biasing the results. The resulting optimized models identified consensus brain regions important for ASD diagnosis consistent across all models. Abnormal functional connectivity (FC) was identified in previously underreported connections to and from the cerebellum and supplementary motor cortex. Furthermore, we characterized the granularity of brain parcellation and feature-set combinations using atlases at differing resolutions to highlight detectable differences in neuroimaging for accurate diagnoses of ASD. Our top performing models match the leading performance in the existing literature, but with an added advantage of lower complexity. This, in turn, makes them more interpretable, less susceptible to overfitting training data, and have a greater ability to generalize to new datasets, which we demonstrate. The generalizability to new data, cross-model consensus, and unbiased optimization all support the validity and robustness of the identified novel connectivity biomarkers. Trustworthy, stable biomarkers validated across multiple models and datasets advance our core neurobiological understanding of ASD, further the promise of machine learning as a diagnostic tool, and best leverage ML’s tremendous potential for accurate, automated medical diagnosis.

## 2 Results

The primary results of this study stem from the analysis of 915 participants from the IMPAC dataset who received both sMRI and resting state functional MRI (rsfMRI)[3]. This study focused on the comparison of two-category classifiers that predict the diagnosis: ASD or Typically Developing (TD). The IMPAC dataset includes an expert clinical diagnosis (the classifier target) for which there were 418 ASD patients and 497 participants designated as TD.

### 2.1 Model performance

The results of our hyperparameter model search for each of our 15 different feature sets and 12 different model types analyzed are summarized in **Fig. 1**, which shows the area under the ROC curve (AUROC) of different machine learning models predicting ASD vs TD on the held-out test data not used during model training. There were 50 hyperparameter configurations evaluated per model by feature set combination, and the feature sets consisted of different brain-atlas parcellations measuring connectivity, volumetry, or both. Each entry in **Fig. 1** shows the AUROC of the model on the test set using the hyperparameter configuration that had the highest average AUROC across the 3 cross-validation folds.

**Figure 1:**
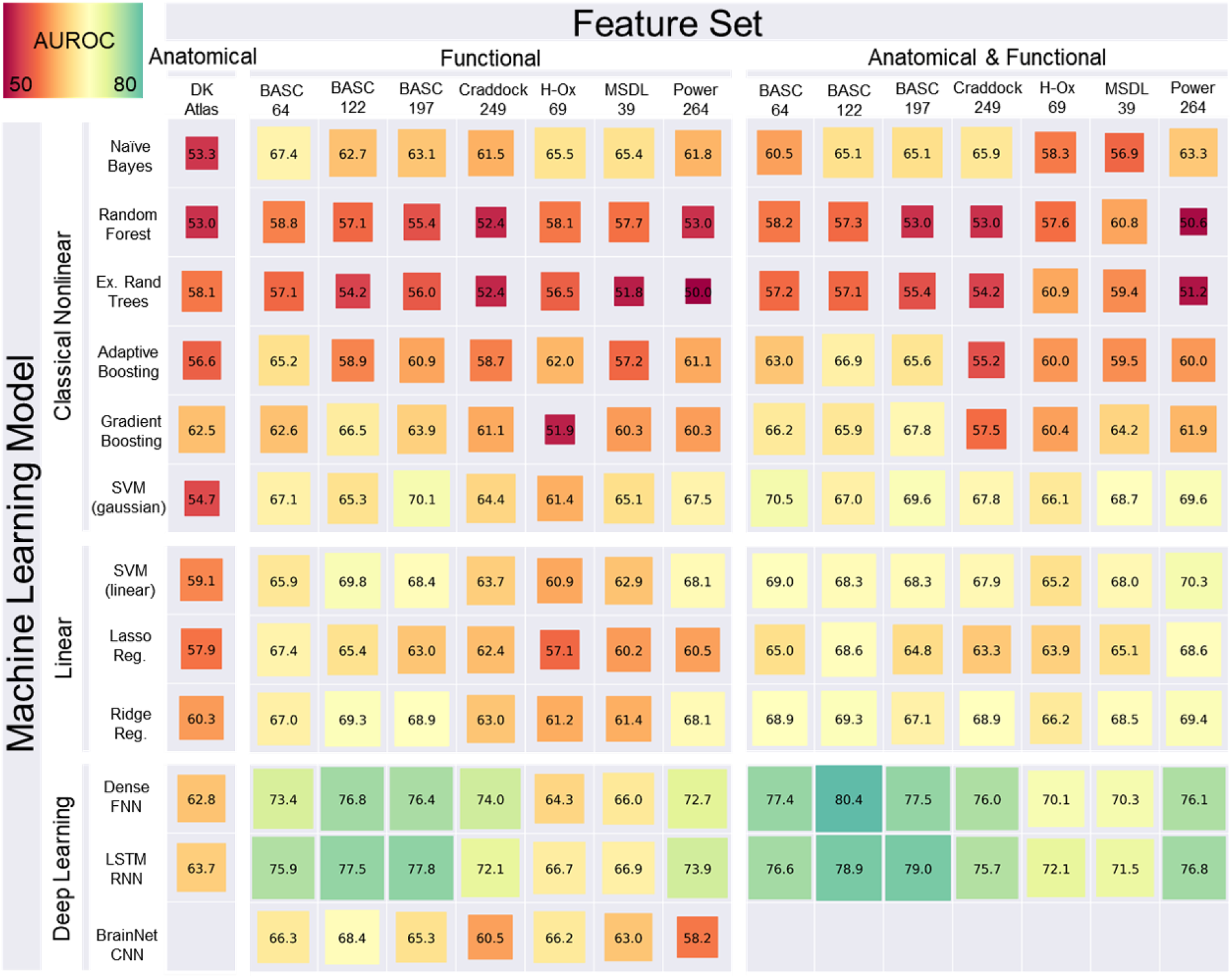
Performance of classifiers predicting the diagnosis of ASD versus TD. Performance is measured as the area under the ROC curve (AUROC) on held-out test data from IMPAC. Greener colors and larger boxes indicate higher performance. Note that BrainNetCNN can only be applied to fMRI data. Rows indicate the type of machine learning model used while columns indicate the feature set (voumetry, connectivity based on a brain-atlas parcellation, or both) used in model training. Models were further broken into Linear, Nonlinear, and deep learning methods. Features were grouped into anatomical, functional, and combined anatomical and functional features. See section 4 (Methods) for further detail.

#### 2.1.1 Impact of machine learning model

The choice of machine learning model category (classical linear, classical nonlinear, and deep learning) had a profound effect on model performance (rows of **Fig. 1**). Deep learning models tended to outperform classical linear models, which in turn tended to outperform classical nonlinear models. The performance of the deep learning models is shown in the bottom 3 rows of **Fig. 1**. The most successful deep learning algorithms were the dense feedforward neural network (DFNN) and long short-term memory network (LSTM), with maximum AUROC of 80.44% and 79.00%, which outperformed classical ML methods. The

BrainNetCNN does not handle anatomical features, however on the functional features alone it performed lower than the other deep learning models and similar to the linear models. The deep learning methods performed best when using the combination of functional and anatomical features. The highest overall performance was a DFNN, whose architecture is described in **Table 1**, right column, using the rsfMRI connectivity data with the BASC atlas with 122 regions-of-interest (ROIs) and the sMRI volumetric data combined, achieving an AUROC of 80.44% on the held-out test set.

**Table 1:** Examples of DFNN network architectures tested in the random search. Hyperparameters shown include the regularization coefficient, number of layers, number of neurons per layer, and dropout fraction. Left column illustrates a simple network with fewer and smaller layers. Middle shows a complex network with more, larger layers. Right column shows the architecture of the highest performing network

| Simple dense network | Complex dense network | Highest performing dense network |
| --- | --- | --- |
| L2 Regularization: 2.3e-4 | L2 Regularization: 2.3e-4 | L2 Regularization: 1.1e-4 |
| Dense 16 neurons | Dense 128 neurons | Dense 64 neurons |
| Dropout: 53% removed | Dropout: 18% removed | Dropout: 13% removed |
| Dense 16 neurons | Dense 128 neurons | Dense 64 neurons |
| Decision Layer 1 neuron | Dropout: 18% removed | Decision Layer 1 neuron |
|  | Dense 64 neurons |  |
|  | Dropout 18% removed |  |
|  | Dense 42 neurons |  |
|  | Decision Layer 1 neuron |  |

Among the linear classical machine learning algorithms, the SVM with a linear kernel and the logistic regression with ridge regularization achieved an AUROC of 70.35% and 69.39% respectively. Among the nonlinear classical machine learning algorithms, the SVM with a Gaussian kernel attained a maximum AUROC of 70.53%. The least successful methods were models from the nonlinear classical model category and include the random forest and extremely randomized trees with a maximum AUROC of 60.75% and 60.90%. Both adaptive boosting and gradient boosting performed better than the random forest models, but overall did not perform as well as the linear methods.

The deep learning models identified non-linear combinations of the functional connectivity and anatomical features which maximize diagnostic accuracy, and that accuracy reached an AUROC of 80% in close agreement with other recent works which require access to whole images. For a confirmatory diagnostic test, high specificity is desirable. With 80% specificity, we achieved a sensitivity of 70% with our top model which is approaching clinical utility (see supplementary **Table S1** for thorough characterization).

#### 2.1.2 Impact of feature set

Upon comparison of the 15 feature sets (columns of **Fig. 1**), we showed that the models trained with only anatomical features (first column) yielded the lowest prediction accuracy. For models trained with functional connectivity data columns (columns 2-8), the BASC atlas and the Power atlas generated models with higher predictive accuracy than other atlases. However, the models trained using a combination of anatomical and functional features (columns 9-15) attained even higher performance, suggesting the information in the functional and anatomical features is complementary. The top performing models combined the anatomical and connectivity features from the Power atlas, Craddock atlas, or BASC atlas. The best performance was achieved with the BASC atlas [4] compared to the other atlases tested. These models achieved 75.4-80.4% AUROC on the held-out test data. This atlas’ coarsest resolution contains 64 ROIs (**Fig. 2A**), its medium-grained granularity has 122 ROIS (**Fig. 2B**), while its fine-grained granularity has 197 ROIs (**Fig. 2C**), and the highest diagnostic prediction performance was achieved at medium granularity. Lone functional features performed higher than structural features, suggesting greater information contained therein. In general, however, combining anatomical features with functional connectivity features tended to improve model performance across all model categories, suggesting structural-functional complementarity. This finding corroborates previous studies [5–11]. The relatively higher performance of the 122- and 197-ROI BASC atlases suggests the optimal granularity of neuroimaging-detectable changes in functional connectivity in ASD. It also indicates that the k-means clustering approach from which the BASC atlas is derived may be more suited to accurately elucidate functional connectivity changes in ASD than other parcellation methods based on anatomical structures.

**Figure 2:**
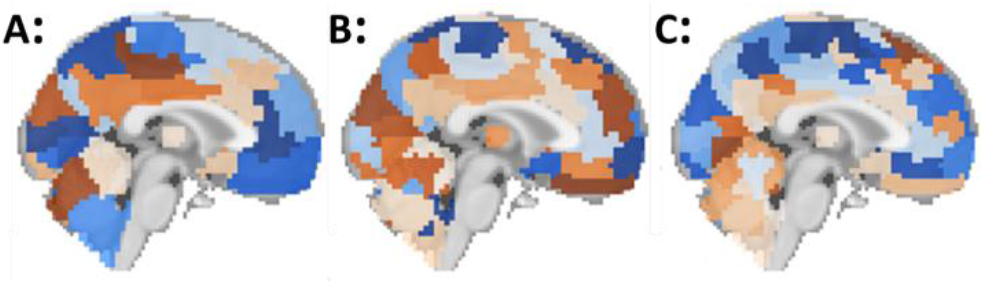
Levels of granularity tested from the BASC atlas. (A) Coarse-grained with 64 ROIs, (B) medium-grained with 122 ROIs, (C) Fine-grained with 197 ROIs.

### 2.2 Important features

As described in section 4.7, the top 15 features were ranked by their median feature importance over the top 5 DFNN models for each BASC atlas (ROI) granularity. These features are shown in **Fig. 3**. The feature importance for the connectivity features are reported as the number of standard deviations from the mean feature importance, i.e., a z-score normalized importance. The most important features for the ASD vs TD prediction for the model trained with 64 ROIs is shown in **Fig. 3A**, while **Fig. 3B** and **Fig. 3C** show the most important features for the models trained from 122 and 197 ROIs, respectively. Color-coded functional labeling of features is shown to facilitate comparison. Motor, sensory, and language areas appear throughout the top features, while no structural features (cortical thickness, volume, etc.) were among the top 15 most discriminative features. Further, whether the connection is significantly increased in ASD (+), decreased in ASD (−), or not significantly different from TD (*o*) is also indicated, as determined by an independent t-test between the ASD and TD sub-cohorts in the study with significance threshold *p* ≤ 0.05. The clinical and demographic features, specifically sex and imaging site, were found to be of high importance only in the 197-ROI model incorporating fMRI and sMRI and models trained on sMRI data alone (not pictured). Additional visualizations of the most important connections in brain space are shown in supplemental **Fig. S1**.

**Figure 3:**
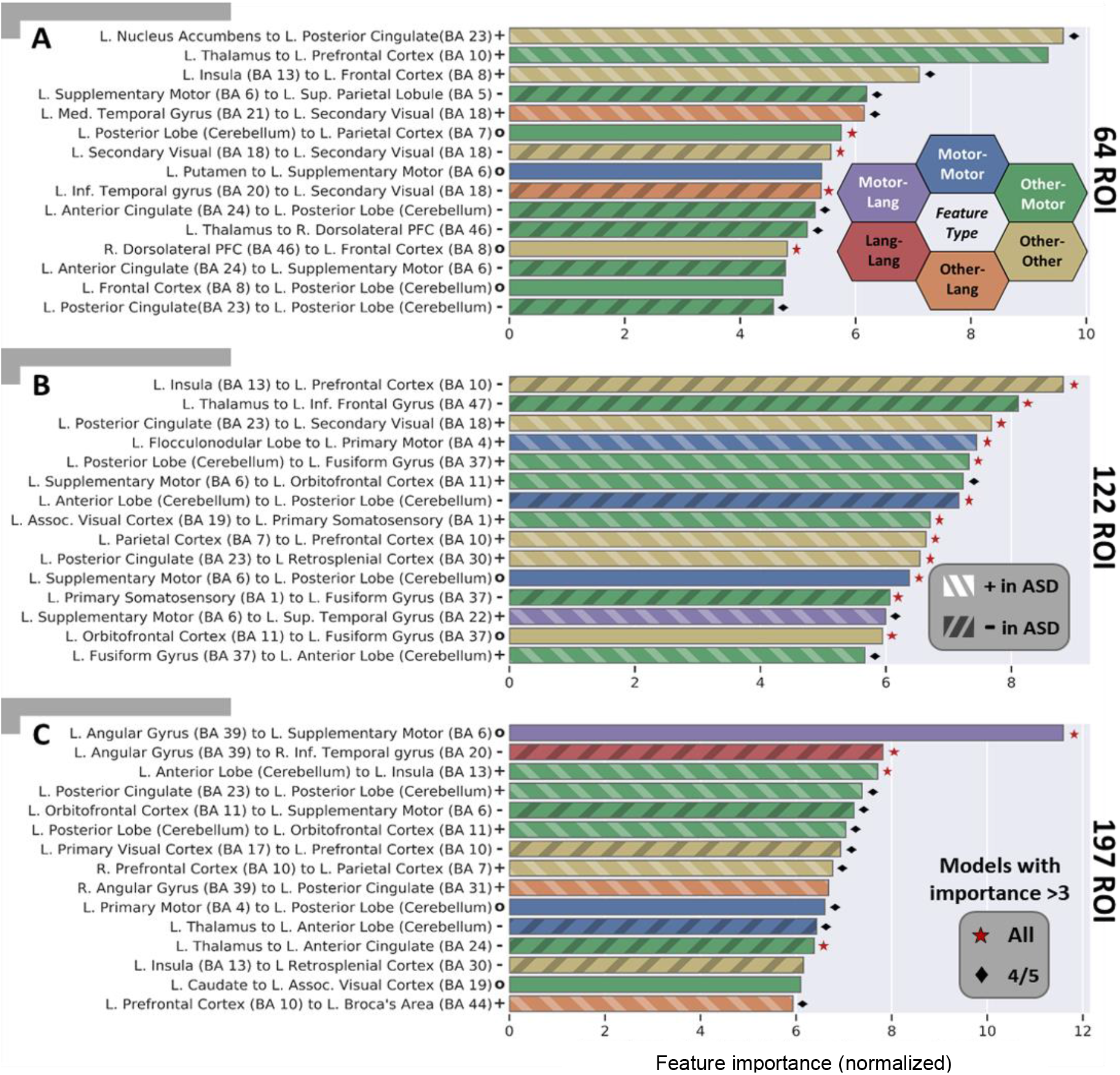
Important features learned by top performing models for ASD diagnosis at each level of BASC atlas (ROI) granularity with coarse-grained atlas (A), medium-grained (B), and fine-grained (C). Each feature captures the functional connectivity between two brain regions and is given a distinct color based on the function of the region pair. Connections between sensorimotor ROIs are shown in blue, while connections between language ROIs are in red. Connections between regions that are neither motor nor language are in yellow. A connection between language (red) and motor (blue) ROIs is shown with an intermediate hue (i.e. purple) and similarly for other region function combinations. For each important feature, an independent sample two tailed t-test testing for a difference in that feature amongst ASD subjects vs. TD subjects was performed. Those that were found to be significant at *p* ≤ 0.05 were additionally marked as increased in ASD (+) and hashed lighter or decreased in ASD (−) and hashed darker. Those connections that were found not to be significantly different in ASD vs. TD were marked with a (*o*) and are presumed to be important features only in a multivariate combination with other features. The corresponding p-values (FDR corrected at 0.01) for the calculated PFI of the median model are significant for all displayed features. This indicates the importances are greater than what would be found by chance due to the random permutations in the PFI approach alone. For each feature, if all 5 of the interrogated models had a z-scored importance ≥ 3, the median feature is marked with a red star, if four had a z-scored importance ≥ 3, the median feature is marked with a black diamond.

Across the different atlas granularities, altered functional connectivity (FC) was found between multiple pairs of brain regions. The *Somatosensory cortex* tended to have altered connectivity (increased and decreased FC) to regions around the brain while the *anterior* and *posterior cerebellum* had decreased FC to deep cortical structures and increased FC to more superficial structures. Meanwhile the *frontal cortex* tended to have a complex pattern of FC changes, *striatal* structures exhibited decreased connectivity with other regions, and *language* associated cortex was found to have a complex pattern of FC changes as well. The default mode network (DMN) encompassed several of these regions, and intra DMN connectivity was significantly altered in ASD vs TD subjects (**Fig. 3**). These patterns of connectivity were found to be consistently important for ASD classification across the atlas granularities examined (**Fig. 3**). Overall, motor associated features were most often predictive of ASD, relative to the other types of features examined. Features recurring at multiple resolutions bolsters confidence in their importance and suggests that even higher granularity may be warranted to further elucidate biological underpinnings.

### 2.3 Model search analysis

Performance of a diagnostic model on a given problem can depend substantially on the choice of architecture. In order to examine the effect of the choice of hyperparameters, kernel density estimates were computed to estimate the probability distribution functions of the configurations of the highest performing (top 20%) configurations and lowest performing (bottom 20%) configurations (**Fig. 4**). As the peaks of the high (blue) and low (orange) performing models are not proximal and the AUROC varied by 20% or more between high and low performing models, this suggests that architectural hyperparameters impact performance substantially. Also, the configurations of the top performing models, i.e. at the peaks in the blue surfaces, occur near the centers of the search ranges and not near the edges of the search space, which confirms that the search ranges used have adequate coverage to discover high-performing configurations. As an additional test, predictive model ensembles combining multiple higher-performing configurations were generated, however these did not further improve prediction performance (supplementary **Fig. S2** and **Table S2**).

**Figure 4:**
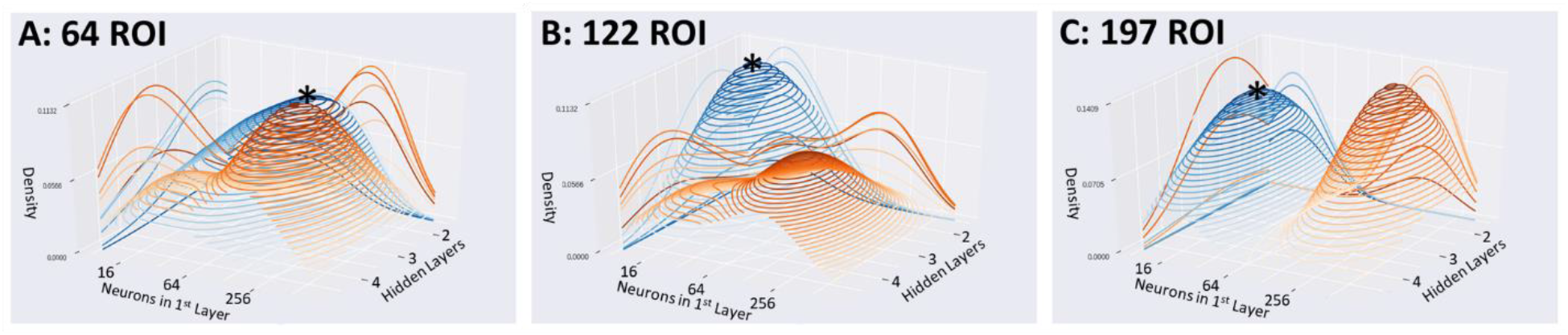
Kernel Density Estimates from the DFNN hyperparameter search. reveals the density of highest performing configurations (top 20%) shown in blue, and low performing configurations (lowest 20%) in orange. Densities of DFNN configurations using the coarse BASC atlas (A), medium atlas (B), and fine atlas (C). Peaks of blue surfaces are marked with *.

The hyperparameter search analysis revealed that the highest performing models tended to use between 2 and 4 hidden layers with 16-64 neurons per layer when using the coarse atlas (**Fig. 4A**), 2 layers with 16-32 neurons versus 3 layers with 128 neurons for the medium-grained atlas (**Fig. 4B**), and 3-4 layers with 16 neurons versus 2 layers with 256 neurons for the fine-grained atlas (**Fig. 4C**). There was a preference in high performing models for more layers with increasing granularity and a preference for fewer neurons/layer with increasing granularity. Models trained on a greater number of features (more granular atlases) tended to perform best with a deeper yet more narrow architecture. Such an architecture would facilitate suppression of spurious features through the narrower ‘information bottleneck’ design, yet still enable the integration of informative features in complex ways through greater architectural depth.

### 2.4 External validation

To test whether the top machine learning models trained with IMPAC captured generalizable predictive abstractions, these models were applied *without adaptation* to two large external datasets (ABIDE I and ABIDE II) not used during model training. Results of this external validation are shown for the ABIDE I and II datasets in **Table 2**. The top single model trained using IMPAC attained 80.4% AUROC on IMPAC held-out test data. When tested on ABIDE I, it achieved an AUROC of 86.0% (**Table 2** underlined) and when tested on the ABIDE II it achieved a performance of 79.2%. These results are very similar to the results attained on the original IMPAC dataset, demonstrating the generalizability of the models and important features identified. A full characterization of other high performing models in this study and their sensitivity-specificity characteristics is presented in supplementary **Table S1**.

**Table 2:** Model performance ranges (AUROC) of the top performing IMPAC models on ABIDE I and II. Highest AUROC of both individual models and highest average AUROC across the top five models are bolded.

| | Atlas | Top 5 models from IMPAC | | | | | Mean ± std |
| --- | --- | --- | --- | --- | --- | --- | --- |
|  |  | 1 | 2 | 3 | 4 | 5 |  |
| ABIDE I | BASC (64 ROIs) | 82.3 | 66.3 | **83.9** | 78.9 | 81.6 | 78.6±6.4 |
| | BASC (122 ROIs) | <u>86.0</u> | 87.1 | **88.0** | 88.0 | 87.4 | 87.3±0.7 |
| | BASC (197 ROIs) | 82.1 | **88.5** | 87.4 | 85.9 | 87.7 | 86.3±2.3 |
| ABIDE II | BASC (64 ROIs) | 74.0 | 64.0 | **77.4** | 73.1 | 74.2 | 72.5±4.5 |
| | BASC (122 ROIs) | <u>79.2</u> | 80.8 | 80.7 | **80.9** | 81.4 | 80.6±0.7 |
| | BASC (197 ROIs) | 76.0 | 82.6 | **82.7** | 79.2 | 82.3 | 80.6±2.6 |

## 3 Discussion

### 3.1 Constructing high performing models

The results of this study yield insight into appropriate mechanisms for the construction of models to inform clinical diagnosis of Autism. The model and atlas characteristics analyzed, when taken as a whole, provide guidance and context for the selection of modeling parameters and features when constructing ASD diagnostic models. *First*, comparison of model types reveals that deep learning provides additional predictive power over classical machine learning methods, even when using engineered image-based features. *Second*, the subset of atlases that performed better is informative for ASD diagnosis. Intermediate granularity atlases generated with functional clustering performed best. *Third*, the results confirm that changes in ASD are reflected more by changes in functional connectivity than by changes in volume and cortical thickness. The results here provide an equitable comparison to guide future neuroimaging experimental design decisions.

### 3.2 Reproducible features

The interrogated models demonstrate important features that are consistent and reproducible across different atlas granularities, the top 5 models within a single granularity, cortical functional types, and with previously published literature. The broad consensus across these domains underscores the credibility of identified biomarkers.

Changes observed in the connectivity between regions in ASD include those at the coarse 64 ROI resolution, where we observed a predominance of decreased connectivity in *sensorimotor* areas in the identified features. Furthermore, we saw prominent involvement of deeper brain structures such as the *cingulate, thalamus*, and *insula*. At the finer resolution of 122 ROIs, we saw a slight shift of the predominantly important features, further involving regions of *association cortex* and a greater number of features with increased FC in ASD versus TD compared to the coarse resolution. Notably, these features are more reproducible than at any other granularity, with feature importances greater than three standard deviations away from the mean in all 5 models or 4/5 models for all of the top 15 median features across the 5 models. This consistency across models bolsters confidence in the 122-ROI models’ features. The features include diverse areas of the brain, but more alterations in FC between regions and the *frontal cortex* specifically are observed. Finally, at the finest resolution of 197 ROIs, we saw many of the same features as at the coarser resolution, with further implication of *association cortex* and *somatomotor* regions. The 197-ROI models did not exhibit a predominance of increased or decreased FC features, unlike the coarser resolutions. Further, at this finest resolution, prolific involvement of deep cortical structures was observed.

Multiple cortical functional networks associated with symptomatology of ASD are implicated at every resolution. The language cortex, corresponding to observed communication differences; the somatosensory processing cortex, corresponding to repetitive behaviors and sensory processing differences; and the social association cortices, linked with social interaction were all observed to have altered connectivity across the models and resolutions observed [1,12–16]. We saw the most FC alterations in both somatomotor and association cortex, with fewer language-associated areas implicated. Many of the features identified by the proposed top performing models agree with alterations reported previously, including the significantly altered DMN connectivity, [13,16,17], connectivity in visual areas [12,15,17,18], motor and supplementary motor connectivity [12], connectivity in somatosensory association areas [14,15], and connectivity in the prefrontal cortex [13,14,16] in individuals with ASD. Importantly, our analysis has better characterized underreported connectivity changes. We showed the FC to and from the cerebellum, including both the anterior and posterior aspects, are important diagnostic predictors of Autism. Cerebellar dysfunction has long been implicated in autism [19–21], but not, to our knowledge, characterized in the multivariate context of machine learning diagnostic models. Moreover, these cerebellar features are important across all levels of granularity examined (from the BASC atlas at 64, 122, and 197 ROIs). These reproducible discriminatory connections lie between the cerebellum and motor areas as well as between the cerebellum and frontal cortex, regions that pertain to sensory processing and social behavior, putatively altered in ASD. This altered cerebellar connectivity in ASD has received little attention in the fMRI literature, as the cerebellum is often not included in functional analyses. We suggest that these connections are areas worthy of further investigation and that all fMRI studies of ASD should especially consider the cerebellum.

### 3.3 Comparison to previous work

This study identified and measured consensus neuroimaging features that were reproducible across processing methods, models, and multiple large datasets. The findings also reproduced previous results from the literature. Improved reproducibility and confidence in experimental results is fundamentally important in the neuroimage analysis community. To maximize reproducibility, it has been shown that performing multiple analyses on the same data and building a consensus from the aggregate results is more reliable than any given single model[22]. For example, when an fMRI model is fit to 212 subjects, the confidence bounds on the estimated performance and held-out test performance are large: greater than ± 15%. The confidence interval follows a binomial law and drops precipitously to ± 2% when the number of subjects is increased towards 1000 subjects [23]. Due to the small effect sizes observed in fMRI studies, the use of multiple datasets and analysis techniques is paramount[24].

Previous research has analyzed the IMPAC [3] and ABIDE [1,2] datasets as well as additional proprietary datasets. This work compares favorably to the top 10 submissions from the IMPAC challenge. The majority of those methods were ensembles of linear models, achieving an average of 0.79±0.01 AUROC [25]. We additionally report AUROC on the external ABIDE I and ABIDE II datasets, confirming that the models generalize (80% AUROC on IMPAC, 86% AUROC on ABIDE I, and 79% AUROC on ABIDE II). There is a scarcity of reported results demonstrating that models trained generalize to other datasets. Our study reports binary accuracy of 75% on the held-out test set. When we trained on IMPAC data and tested the model on ABIDE I and II, our model’s test accuracy is comparable to models trained *directly on ABIDE data*. For example, models using training data sampled across all ABIDE sites report test accuracies ranging from 64-68% [26,27], while those holding out whole sites [8,25,28] report test accuracies from 72-80%.

Furthermore, often results on the ABIDE dataset report accuracy on the same validation data used to optimize model hyperparameters, rather than separate test data, which tends to overestimate classifier performance [23]. These previous reports include those using classical machine learning, typically ensembles [5,9,11,28–35] with validation accuracy from 65-83%, and those using deep learning [9,31,35– 38] with validation accuracy 70-85%. Finally, previous research analyzing proprietary datasets [9,10] achieve validation accuracy of 78-92%. In contrast to this prior work that use private inaccessible data or proprietary modeling, this work employs large publicly available datasets and the code is publicly available through our source-code repository (see section 6: Data availability). Such steps foster greater potential for external reproducibility and verification.

In summary, this study addresses gaps in the aforementioned research: First, we perform extensive quantitative comparison across multiple model types and atlas granularity, where most studies analyze only a small subset of models and atlases. Second, we verify model performance on a large, public, external dataset and demonstrate generalizability, which to our knowledge, has not been done before. Third, we report *test* performance on both the IMPAC and ABIDE datasets because validation accuracy overestimates model performance. Finally, since there is little discussion of the consistency of important features across models, across atlases, and atlas granularity, we report those features which are consistently important.

### 3.4 Limitations and future directions

While our study significantly advances the development of machine learning tools for automated accurate ASD diagnoses, it has potential for improvement. The model is dependent upon the input dataset, and the IMPAC dataset has only binary diagnosis. However, ASD is known to be a spectrum disorder. Training data that includes a finer characterization of ASD symptomatology would help hone the accuracy and enable a fuller characterization of the disorder. Additionally, our analysis used only one anatomical parcellation, but additional structural atlases could be explored to provide a better integration with the functional connectivity. Finally, data that includes measures of electroencephalography (EEG) and magnetoencephalography (MEG) which directly measure brain activity albeit at lower special resolution than fMRI would complement our analyses. Future studies to explore the character of ASD as a spectrum, integrate additional functional and anatomical measures, and explore different timescale resolutions would further advance our understanding.

### 3.5 Conclusion

This study systematically compares 12 of the most powerful and commonly deployed ML models, develops a high performing ASD diagnostic model that can be readily adapted to new datasets, and characterizes the important and reproducible features learned by the models. Predictive features learned by the models confirm previously reported putative biomarkers and place new importance upon the understudied *in-vivo* connectivity between the cerebellum and the supplementary motor and frontal cortices. The identification of optimal brain parcellation granularity and feature-set combinations can be used to further guide model development, develop clinical diagnostics, and improve ASD diagnosis and timeliness of care. The identified putative biomarkers may help to elucidate pathophysiology, direct treatment options, and even target psychosocial interventions. Building evidence and confidence in identified neurophysiologic correlates of autism will benefit the community and individuals affected.

## 4 Methods

### 4.1 Materials and Ethics statement

This study uses the 915 participants of the IMPAC dataset that received both sMRI and resting state functional MRI (rsfMRI)[3]. This dataset includes an expert clinical diagnosis (the classifier target) for which there were 418 ASD patients and 497 participants designated as TD. Demographic data including participant age and sex were collected (**Table 3**). To test whether the machine learning models trained with the IMPAC dataset captured discriminative features that generalize to other data, two external ASD datasets with sMRI and rsfMRI were used, ABIDE I [1] and ABIDE II[2]. Demographics for the participants used from ABIDE I and ABIDE II are shown in **Table 3**. Participants from all sites of the ABIDE I and ABIDE II were included for external validation, provided both sMRI and rsfMRI were obtained on the same visit. The first available pair of sMRI and rsfMRI scans were used per subject. 1045 subjects from ABIDE I and 761 subjects from ABIDE II met this criteria.

**Table 3:** Demographics of IMPAC participants, ABIDE I participants, and ABIDE II participants. Errors shown are standard deviations.

|  | IMPAC | ABIDE I | ABIDE II |
| --- | --- | --- | --- |
| *Female %* | 50 | 15 | 20 |
| *ASD %* | 46 | 48 | 52 |
| *Age (years)* | 17±9.6 | 17±8.1 | 17±11 |
| *Total Participants* | 915 | 1045 | 761 |

The IMPAC data used for the analysis in this study were anonymized with no protected health information included and was approved by the ethics committees of the Institut Pasteur, Robert Debre Hospital, Paris-Saclay Center for Data Science, and Ingenieurs et Scientifiques de France [3]. The ADNI data used for analysis in this study were anonymized with no protected health information included in accordance with NIH guidelines and HIPPA guidelines and conform to the ethics standards set in the 1000 Functional Connectomes Project and INDI [1,2]. All data was gathered with informed consent from all participants.

### 4.2 MRI feature extraction

The IMPAC fMRI and sMRI were processed using the fconn1000 pipeline [39]. For the fMRI connectivity features, the TSE connectivity metric was fit on either the training data alone (IMPAC models) or the entire IMPAC dataset (ABIDE 1 and 2 datasets), and the TSE values per-ROI timeseries for the ABIDE dataset were calculated. Identical structural and functional features were derived as described above for the IMPAC study using FreeSurfer volumetry and tangent space embedding between mean regional timeseries (with the originally calculated IMPAC embedding) respectively [40,41]. The features were standard scaled by the mean and standard deviation of the IMPAC dataset. Further details of the preprocessing are provided in supplemental section 1.1.

A schematic of the feature extraction procedure is provided in **Fig. 5**. As there is no atlas which is optimum for every prediction task, multiple independent atlas parcellations were used. From the rsfMRI, functional connectivity matrices were derived as illustrated in **Fig. 5A,B**.The rsfMRI was first parcellated into regions of interest (ROIs) using seven different atlases. The first three atlases (1-3) are variations of the BASC atlas. This regions of this atlas are defined by k-means clustering of stable coherent groups [4] and 3 atlas granularity levels are tested with 64, 122, and 197 ROIs. The fourth atlas is the Craddock atlas, which defines 249 ROIs by coherence of local graph connectivity [42]. The fifth atlas is the Harvard-Oxford Anatomical atlas, which defines 69 ROIs using anatomical features. The sixth atlas is the MSDL atlas, which has 39 ROIs defined by correlations of spontaneous activity [41]. The seventh atlas is the Power atlas [43], which divides the brain into 264 ROIs based on local graph-connectivity. The mean rsfMRI time signals from each ROI were converted into a connectivity matrix by projection into tangent space, which better captures subject-specific variations from one or more groups than correlation alone [44]. The tangent space embedding (TSE) projection was fit on the training data alone and applied to the test set.

**Figure 5:**
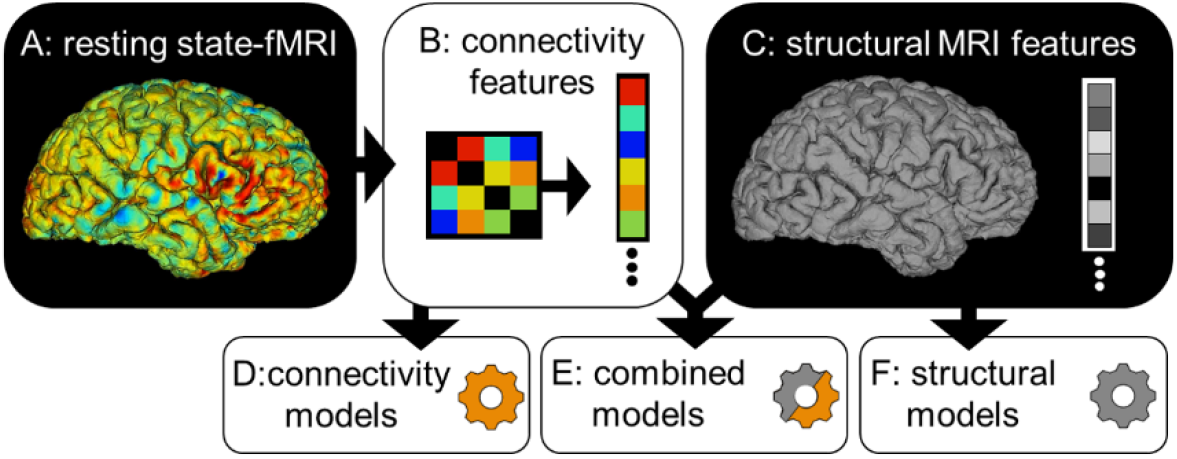
Combinations of derived features used by the predictive models evaluated in this study. (A) The rsfMRI was transformed into a symmetric connectivity matrix for each atlas. (B) Upper triangular elements of matrix were flattened into a 1D vector. (C) The sMRI was transformed into a vector of cortical and subcortical ROI volumes and cortical thickness features. Different combinations of fMRI and sMRI features were compared: In (D) the connectivity matrix vector is used as the sole input for the predictive model, in (E) both anatomical and connectivity derived feature vectors are concatenated and used, while in (F) the anatomical features are used as the sole input for the predictive model.

From the sMRI, 207 features were extracted with Freesurfer 6.0, including volumes of 68 cortical and 37 subcortical structures, as well as regional cortical thickness and area for the ROIs defined by the Desikan-Killiany gyral atlas [45]. The extraction of this anatomical feature vector is schematized in **Fig. 5C**. Then, models were fit using either the structural features (**Fig. 5D**), functional features (**Fig. 5F**) or both (**Fig. 5E**).

### 4.3 Data partitioning

IMPAC participants were randomly partitioned with 80% assigned to a training set and 20% to a test set with the splits having matching proportions of diagnosis (ASD/TD) and sex (male/female). The test participants were set aside and not used during training or model selection. The training set was further split into validation and training folds using a 3-fold stratified cross validation approach. To ensure fair subsequent model comparison, the same splits were used for all tested machine learning models.

### 4.4 The machine learning models

Systematic testing of a broad array of 12 machine learning classifiers was conducted. These models were chosen to span statistical complexity and to be representative of models with evidence of high performance in previous ASD studies. We used 3 linear classical ML models with lower statistical complexity [9,26,30,33,34,46], 6 non-linear classic ML methods of moderate statistical complexity [8,10,11,26,29,32,34,46], and 3 deep learning approaches with higher statistical complexity [26,28,37,38,46,47]. These models are listed with their hyperparameters in **Table 4**. Classical models were constructed using the Scikit-learn and XGBoost packages, while the deep learning models were implemented with Keras, Tensorflow, and Caffe packages [48–52]. The LSTM classifier uses a dense neural network atop a bidirectional LSTM for classification as in [53]. This has been shown to yield high prediction performance even on non-sequential fixed vector data [54]. The graph-convolutional network classifier, BrainNetCNN, was trained using just the FC matrix [55].

**Table 4:**
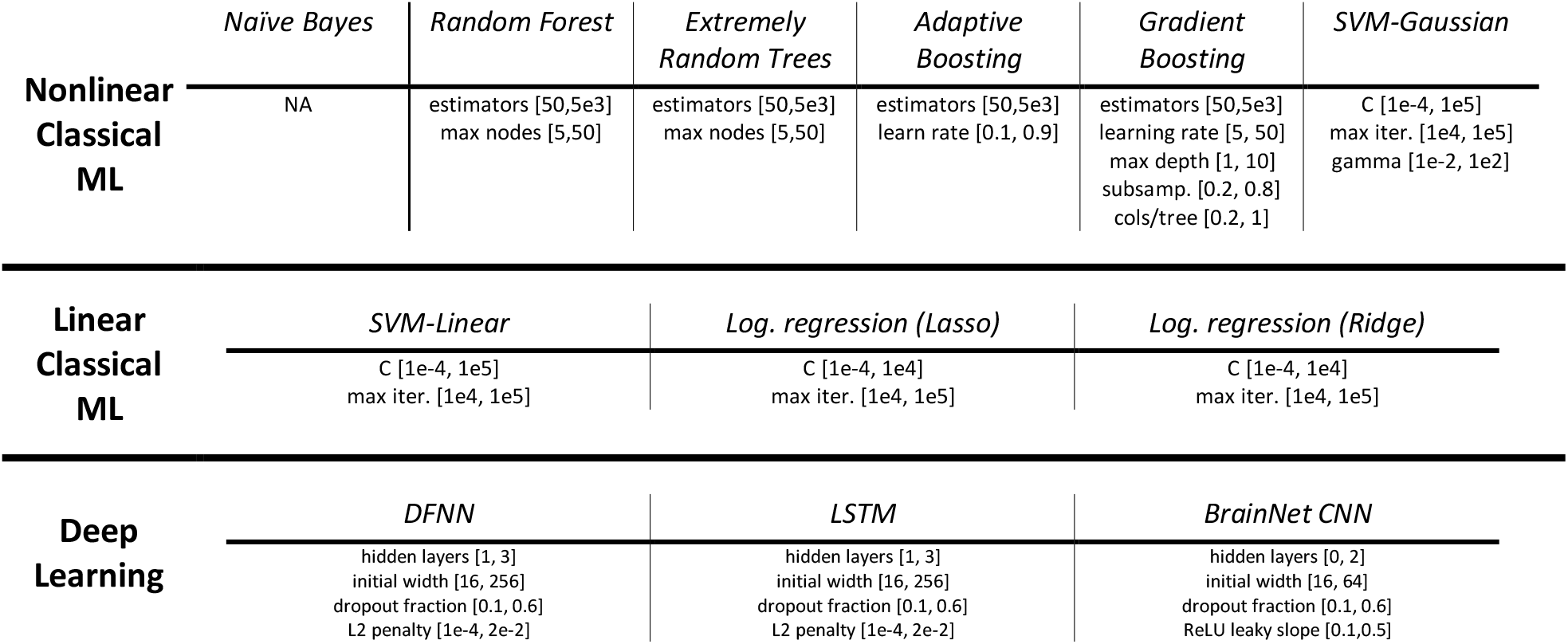
Hyperparameter ranges for each machine learning model. Abbreviation definitions (in order of appearance from left to right): ML=Machine learning, learn rate = learning rate, subsamp. = subsampling per tree, cols/tree = columns per tree, max iter. = maximum iterations, SVM = support vector machine, SVM-Gaussian = SVM with gaussian radial basis function kernel, SVM-Linear = SVM with linear kernel, Log. regression = logistic regression, lasso = lasso regression with L1 penalization, ridge = ridge regression with L2 regularization, DFNN = deep feedforward neural network, LSTM = bidirectional long short term memory neural network, BrainNet CNN = BrainNet convolutional network [52], ReLU leaky slope = rectified linear activation unit slope for x<0.

### 4.5 Training the models

Each of our 12 model types was trained on 15 different feature sets, for a total of 180 model type by feature set combinations. The feature sets contain measures of anatomical volume and functional connectivity from the IMPAC dataset. These feature sets included: (1-7) functional connectivity measured between regions defined by one of the 7 atlases described in section 4.2 (using the processing steps in **Fig. 5A,B,D**), (8) an anatomical feature set consisting of 207 measures of regional volume and thickness (**Fig. 5C,F**), (9-15) the union of the anatomical feature set with one of the functional feature sets (**Fig. 5A-C,E**). All feature sets also included sex and imaging site as additional covariates. The deep learning models were trained on an NVIDIA Tesla p100. Further description of the training of the deep learning models can be found in supplemental section 1.2.

### 4.6 Optimizing model hyperparameters and model selection

In order to achieve good performance, model *parameters* (i.e. weights) must fit the training data and model *hyperparameters* that govern overall characteristics, such as neural network architecture or regularization terms in a regression model, need to be selected. In this study, to ensure fairness across model types, the random search algorithm was employed to provide an unbiased tuning of model hyperparameters, rather than manually tuning which is biased to the developer’s level of expertise. A random search has been found to be more effective than a grid search across a wide variety of model types and inputs due to more samples being taken across highly important hyperparameters [56]. The dimensions and ranges of the hyperparameters searched for each model are listed in **Table 4**. For each model, 50 configurations were randomly drawn from the hyperparameter space. To further ensure fairness, the same data partitioning splits were used for the 3-fold cross-validation partitioning of the training set. For each model category, the highest performing configuration was selected by mean AUROC across the cross-validation folds. The model using this configuration was then trained on all training data and evaluated on the held-out test set, not used in training.

### 4.7 Identifying important and reproducible features

In order to better understand which features were consistently important in the diagnostic ASD/TD prediction, multiple top performing models were further analyzed. In this study, the top model category ranked according to validation AUROC was the DFNN. Its best performing model configurations were trained on the BASC atlas, whose rankings are detailed in results section 2.1. Because they had the highest performance across multiple scales, these models and scales were the subject of further analysis and interrogation to determine their learned features. In particular, for each BASC scale, the top 5 models per BASC atlas parcellation granularity were identified and the top 15 features were ranked by their median feature importance over the top 5 DFNN models (**Fig. 3**).

The importance of each feature for each of these models was computed using permutation feature importance (PFI) [57]. PFI was chosen because it can be applied uniformly to all of the model feature type combinations tested. In this approach, for a given trained model, each feature is individually permuted across all participants to ablate any predictive information present. Its feature importance, *I*, is calculated as the z-score normalized mean decrease in AUROC: *I* = *AUROC*_*b*_ − *AUROC*_*a*_, between the performance before feature permutation (*AUROC*_*b*_) minus the performance after feature permutation (*AUROC*_*a*_). This was averaged over 64 random permutation repetitions. The distribution of calculated importances across the permutations in the median model was compared with a one-tailed t-test to a null distribution of feature importance created following the procedure in [58]. The corresponding p-value was then FDR corrected at a rate of 1% with the Benjamini-Yekutieli procedure [59]. This tests if the importances are greater than what would be found by chance due to the random permutations in the PFI approach alone and if the number of permutations was sufficient to find important features. To aid in the comparison of IMPAC connectivity features to the scientific literature, which often reports results in Brodmann areas (BA), the centroid of each ROI of each atlas was calculated and matched to the corresponding BA [60]. The ROI-ROI connection can then be re-written as the closest BA-BA connection and the corresponding functions compared.

### 4.8 Model search analysis

A hyperparameter search generates a wealth of information. To obtain insights from this information, kernel density estimates were computed for the models with the top 20% of performance and for the models with the lowest 20% of performance across the 3 BASC atlas resolutions (3 of the highest performing atlases) to identify regions of hyperparameter space that tended to distinguish high performing models from low performing ones, as shown in **Fig. 4**.

### 4.9 External validation

Any given model may overfit to spurious information in training data, not capturing the most biologically relevant information, but fitting to noise. A true biomarker should not only be identifiable in multiple models fitted to the same data, but also be predictive when used in an entirely new dataset. To test whether the machine learning models trained with the IMPAC dataset have truly captured discriminative features, we use external datasets (ABIDE I and ABIDE II) not used during model training and hyperparameter optimization. Each of the top 5 DFNN models which used the combined structural features and functional features from the BASC atlas at three resolutions (64, 122, and 197 ROIs) was applied without adaptation directly to the connectivity and anatomical features derived from the external datasets, ABIDE I and ABIDE II.

## Data Availability

To facilitate reuse and extension, we are pleased to provide full source code for the proposed approach at:https://gitfront.io/r/DeepLearningForPrecisionHealthLab/01db3a7959b24dc387a3ae5c824124962a093373/ConsistentASDCorrelates/
Datasets used for analysis during the study are available in the IMPAC repository (https://paris-saclay-cds.github.io/autism_challenge/), and ABIDE I and II repositories (https://fcon_1000.projects.nitrc.org/ indi/abide/)

https://fcon_1000.projects.nitrc.org/indi/abide/

## 5 Acknowledgements

Special thanks to Dr. Michael D. Rugg, PhD, Dr. Satwik Rajaram, PhD, and Dr. Prapti Modi, PhD for providing additional feedback and editing during the writing of this manuscript.

Cooper Mellema was supported by NIH NINDS F31 fellowship NS115348. Alex Treacher and Albert Montillo were supported by NIH NIA R01AG059288. Albert Montillo was additionally supported by NIH NCI U01 CA207091, the King Foundation, and the Lyda Hill Foundation.

## 6 Data availability

To facilitate reuse and extension, we are pleased to provide full source code for the proposed approach at:https://gitfront.io/r/DeepLearningForPrecisionHealthLab/01db3a7959b24dc387a3ae5c824124962a093373/ConsistentASDCorrelates/

Datasets used for analysis during the study are available in the IMPAC repository (https://paris-saclay-cds.github.io/autism_challenge/), and ABIDE I and II repositories (https://fcon_1000.projects.nitrc.org/indi/abide/) [1–3].

## 7 Author contributions

C.J.M. conducted the data processing, model construction, and model interrogation and analysis with supervision from A.M.. K.P.N. and A.T. contributed to data processing and model interrogation. C.J.M and A.M. wrote the manuscript with input from K.P.N., and A.T..

## 8 Competing interests

The authors declare no competing interests.

## Notes

### Competing Interest Statement

The authors have declared no competing interest.

### Author Declarations

Datasets used for analysis during the study are available in the IMPAC repository (https://paris-saclay-cds.github.io/autism_challenge/), and ABIDE I and II repositories (https://fcon_1000.projects.nitrc.org/ indi/abide/)

