## Supplement for "Reproducible neuroimaging features for diagnosis of Autism spectrum disorder with machine learning"

### Supplemental information

#### 1.1 Preprocessing pipeline

For the IMPAC and ABIDE fMRI data, TR per subject varied from 0.8-3 s. Each functional scan was processed by the fconn1000 pipeline [39], where the following steps were performed in order: dropping the first frame, deobliquing, reorienting, motion correction, skull stripping, spatial smoothing with 6mm gaussian filter, grand mean scaling, band pass filtering [0.005-0.1Hz], linear and quadratic detrending, and brain mask generation. Then, the fMRI image was registered to the T1 image and from there to MNI 152 space [61,62]. Finally, potential nuisance signals (global, WM, CSF, and motion parameters) were regressed out.

The structural T1 images were processed in the following order, also by the fconn1000 pipeline [39]: deobliquing, reorienting, skullstripping, registration to MNI 152 space [61,62], and segmentation of brain from background, CSF, and WM segmentation.

#### 1.2 Training of the deep learning models

The deep learning models used the leaky ReLU activation function, early stopping with a patience of 20 and minimum change of 1%, the Nesterov ADAM optimizer, a batch size of 128, and the binary cross-entropy loss function. For both the dense and LSTM networks, the first two layers used the same initial layer width; subsequent layers were each reduced in width by a factor of 2.

#### 1.3 Neuroanatomical locations of important connectivity features

In order to further characterize the most important features for diagnosis learned by the top 5 performing DFNN models, glass brain views are constructed to reveal the anatomical location of the features. These views are shown in Fig. S1 and have a normalized feature importance z-score>=5. This analysis is performed at each of 3 granularity levels of the BASC atlas. Substantial overlap of feature importances across the different granularities tested is observed in terms of brain-regions involved, which is detailed in the sections 4.1.2 and 4.2.

#### 1.4 Further characterizing model performance

The models in this study were trained with a loss function that maximizes AUROC. To further characterize the performance of the top models and to facilitate comparison to the related literature, (e.g., as discussed in 5.3), the accuracy and sensitivity of the high performing models across several specificities has been computed and is shown in **Table S1**.

#### 1.5 Performance using multiple atlases simultaneously

Since it could be the case that multiple atlases provide substantial complementary information, an additional experiment was conducted in which the features derived from pairs of atlases were used as the input to a DFNN model.

**Table S1:** Additional predictive performance metrics. The area under the ROC curve (AUROC) is shown for the top model alone and compared to the performances of the top 5 models using each parcellation. Additionally, base accuracy and sensitivity characteristics for fixed values of specificity are also displayed.

| | | | **Sensitivity at:** | | |
|---|---|---|---|---|---|
| | **AUROC** | **Accuracy** | ***60% Spec.*** | ***70% Spec.*** | ***80% Spec.*** |
| *Top model alone* | 80 | 75 | 77 | 74 | 70 |
| *BASC 64 top 5 models* | 75-78 | 69-71 | 74-80 | 65-71 | 51-62 |
| *BASC 122 Top 5 models* | 76-80 | 67-75 | 77-81 | 68-76 | 57-70 |
| *BASC 197 Top 5 models* | 76-78 | 70-75 | 75-81 | 68-74 | 60-63 |

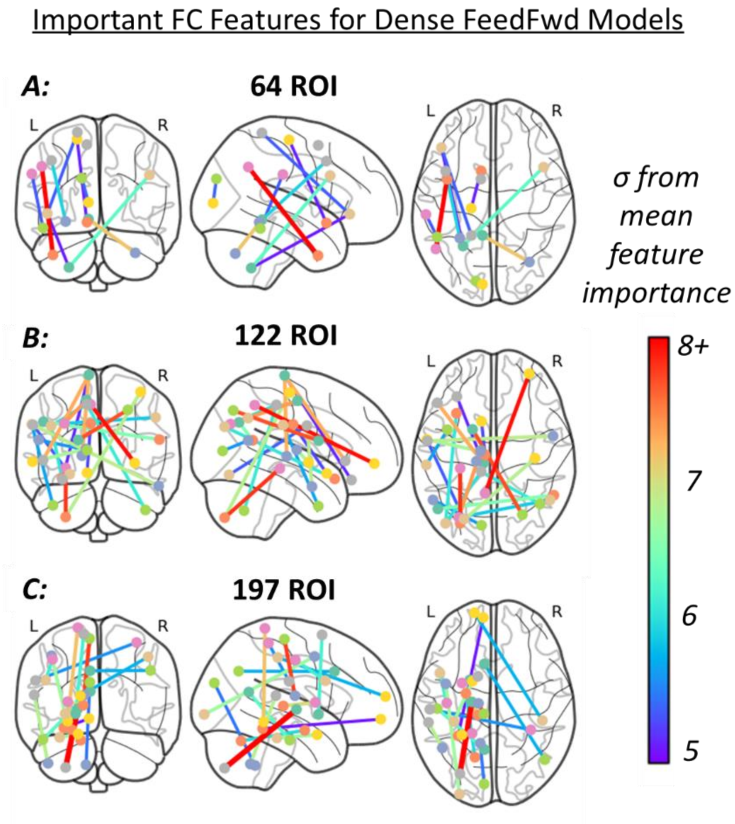


**Figure S1:** Neuroanatomical locations of the most important functional connectivity features and their relative importance. Features for the DFFN model using the BASC atlas with coarse (A), medium (B), and fine (C) granularities. Features with z score >=5 are shown while their color indicates the number of standard deviations they are from the mean feature importance. Node color is for visualization only.

Model performance in these experiments is presented in **Fig. S2**. The models using feature from two functional atlases, or two functional atlases plus the structural (anatomical) features performed no better than models using features from a single functional and the anatomical features. Due to the additional complexity at no benefit to performance, larger combinations were not explored.

#### 1.6 **Ensembled learning models**

Previous reports have suggested that ensemble models may be optimal. The unbiased architecture search, provides ample opportunities to ensemble models to test for improved performance. Therefore, ensembles of models from high performing models were generated as well as ensembles that include new models using combinations of features.

Ten of the highest performing models using disparate atlas by feature set combinations were themselves combined exhaustively in groups of 3 to 5 model ensembles. To determine a diagnosis from the ensemble, soft voting, hard voting, a logistic ridge regression ensemble, and a linear SVM ensemble were all tested exhaustively. The chosen models to combine included: (1) the top 10 performing models of the DFNN network trained on the combined structural and functional MRI parcellated with the BASC atlas with 122 ROIs; (2) the top 10 DFNN's trained on structural and functional data from the BASC atlas with 197 ROIs (3) the top 10 LSTM's trained using the fMRI data alone parcellated with the BASC atlas with 122 ROIs (4)

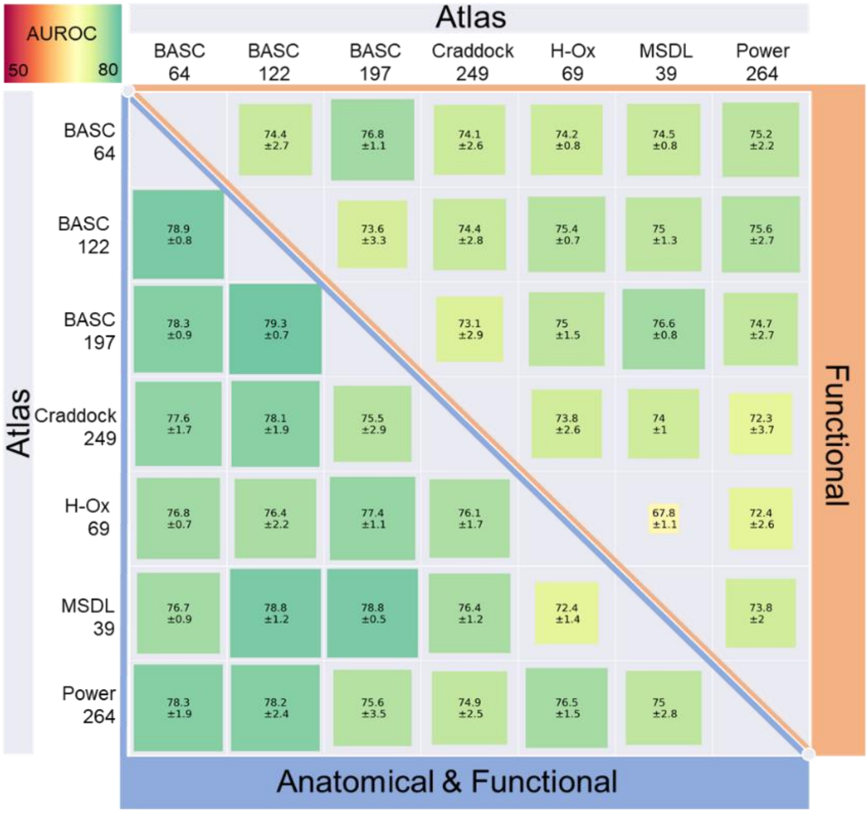


**Figure S2:** Comparing performance of two atlas combinations used as inputs to a DFNN model. The lower triangle outlined in blue also included structural features, while the orange box had only the combined 2-atlas fMRI feature sets (e.g., no cortical thicknesses as input). The error ranges were created with repeated reinitialization and training on the same training data. No combination outperformed the top models using a single functional atlas.

the top 10 LSTM's trained using both the structural and fMRI data parcellated with the BASC atlas with 122 ROIs (4) the top 10 linear ridge regression models using combined structural and functional data and the BASC 122 atlas. This subset was chosen as these 5 categories were high performing models and spanned a variety of machine learning methods and features which may have learned complementary model abstractions for diagnosis. **Table S2** shows the performance of the top ensembles on the held-out test data. The top ensembles were chosen with 10x cross validation on the training set. There were no gains in performance observed over using individual models. Due to the large increase in model complexity, ensembles of even greater complexity were not explored.

**Table S2**: Performance of the top ensembles on the held-out test data. The top ensembles were chosen with 10x cross validation on the training set, and their AUROC on the test set is displayed here.

| | **Soft Voting** | **Hard Voting** | **Logistic Ridge** | **Linear SVM** |
|---|---|---|---|---|
| *Top Model alone* | 0.804 | | | |
| *Ensemble of 3* | 0.77609 | 0.76064 | 0.77730 | 0.75986 |
| *Ensemble of 5* | 0.77297 | 0.74681 | 0.75060 | 0.75565 |
